# The Kinetics of COVID-19 Vaccine Response in a Community Vaccinated Population

**DOI:** 10.1101/2021.09.18.21263605

**Authors:** Michael Tu, Samantha Chiang, Richard Bender, David T.W. Wong, Charles Strom

## Abstract

We used a noninvasive electrochemical quantitative assay for IgG antibodies to SARS-CoV-2 S1 in saliva to investigate the kinetics of antibody response in a community-based population who had received either the Pfizer or Moderna mRNA-based vaccines. Samples were received from a total of 97 individuals including a subset of 42 individuals who collected samples twice-weekly for 3 months or longer. In all, 840 samples were collected and analyzed. In all individuals, salivary antibody levels rose sharply in the 2-week period following their second vaccination, with peak antibody levels being at 10-20 days post-vaccination. We observed that 20%, 10% and 2.4% of individuals providing serial samples had a 90%, 95%, and 99% drop respectively from peak levels during the duration of monitoring and two patients fell to pre-vaccination levels (5%). The use of non-invasive quantitative salivary antibody measurement can allow widespread, cost-effective monitoring of vaccine response.

**Article Summary Line:** COVID-19 antibodies were measured in saliva and 20% of vaccinated subjects experienced a 90% drop in peak antibody levels over the course of monitoring.

## INTRODUCTION

The pandemic caused by SARS-CoV-2 has led to worldwide fatalities and social and economic disruption. In the autumn of 2020, the FDA issued emergency use authorization for two mRNA-based vaccines manufactured by Pfizer/BioNTech COMIRNATY^®^ (Pfizer) or Moderna/NIAID (Moderna). Both vaccines use mRNA sequences from the S1 domain of the SARS-CoV-2 Spike Protein *(1-5)*, and vaccines require two doses given 21 or 28 days apart in order to achieve 95% protection against SARS-CoV-2 infection (*1-5*). It is unclear whether all individuals developed antibodies with 5% at risk of breakthrough infection, or whether a modest fraction of individuals will not respond develop antibodies and remain at risk of infections.

Unlike the predicted statistics for the healthy general population, it is known that patients on immunosuppressive drugs and cancer patients may not develop a robust antibody response to vaccine administration *(7)*. It is possible that a fraction of individuals in the population may have an undetected immune deficiency that prevents them from responding appropriately to the standard vaccine regimen. Consequently, the Centers for Disease Control and Prevention (CDC) is currently recommending booster immunizations be deployed beginning in the fall of 2021*(8)*.

Several studies have demonstrated that circulating antibody levels decrease over time following either vaccination or infection *(9-14)*. Breakthrough infections are being observed in fully vaccinated individuals. It is not known what level, if any, of circulating antibody is required to have immunoprotection against COVID-19 infection. Current publications report very little information regarding the kinetics of antibody levels in patients following vaccination and these studies only report antibody levels at 5.5 weeks and 90 day intervals post second vaccination respectively *(15,16)*. Current research is underway to determine whether the efficacy of booster immunization doses and its timing in protecting against SARS-CoV-2 infection, especially in light of the emergence of highly contagious variants such as the delta variant that may be less sensitive to the current vaccines.

It is clear that most, if not all, individuals receiving both doses of either the Pfizer or Moderna vaccine respond with a robust IgG response *(1-5)*. However, what is lacking, is frequent kinetic monitoring and long-term monitoring of antibody levels in a community vaccinated population. Non-invasive monitoring using saliva allows for frequent and long-term monitoring of vaccinated individuals and entire populations.

We have developed a saliva-based quantitative assay for IgG antibodies to the S1 domain of Spike protein in SARS-CoV-2 using a novel electrochemical platform formerly known as EFIRM^®^ and now called Amperial^®^ *(17)*. Previously, we used this assay to monitor patients who had recovered from COVID-19. This assay was greater than 98% specific for individuals with prior COVID-19 infections and gave proportional results to serum assays performed at the same time on the same patient. Two other groups have similarly demonstrated the ability of saliva to be a surrogate for serum or plasma measurement of SARS-CoV-2 antibodies *(18,19)*.

## METHOD

### SARS-CoV-2 Salivary Assay Equipment

The Amperial^®^ platform uses a proprietary 96-well microtiter plate containing gold electrodes at the bottom of each well and an electrochemical reader system (EZLife Bio Inc, Los Angeles, CA). The description of the Elzie Amperial^®^ COVID-19 Antibody assay and the assay performance and validation have been described previously *(17)* and is summarized in the following section.

### Immobilization of SARS-CoV-2 on Plate Surface

For the preparation of the antigen coated wells we prepare a 10 μg/mL SARS-CoV-2 S1 antigen (SinoBiological US Inc, Wayne, PA) diluted in a solution of 72.25 mM pyrrole (Sigma-Aldrich, St. Louis, MO) and 0.147M KCl mixture. This antigen-polymer mixture was then briefly vortexed for 1 sec and then 30 μL was added to each well. Each plate contains wells containing antigen alternated with wells containing polymer without antigen added. The plate was then inserted into the electrochemical plate device and a square wave potential applied that consists of 1 second of +350 mV and 1 second for +1100mV for 4 cycles (8 seconds total) to electropolymerize the polymer and antigen-polymer on the surface. After the electrochemical polymerization, each electrode was washed for 3 cycles in a buffer of 1x phosphate-buffered saline (PBS, Affymetrix, USA) and 0.05% Tween 20 (BioRad, USA), referred to as PBS-T.

### Sample Preparation and Incubation

Saliva samples were diluted 1:10 in a 1% (w/v) Casein/PBS solution (ThermoFisher, Waltham, MA). Internal standards consist of SARS-CoV-2 IgG antibodies (Absolute Antibody, Oxford, United Kingdom) diluted in 1% (w/v) Casein/PBS solution at varying concentrations in the linear range of the assay to provide a standard curve. Thirty microliters of the samples and standard are then added to their respective wells in the coated electrode plate. All patient samples were added to both a pyrrole only well and an S1 antigen coated well. The plate is incubated at room temperature for 10 min before washing with PBS-T.

### Secondary Antibody and Reporter Enzyme

Subsequently biotinylated Goat Anti-Human IgG Fc (ThermoFisher, Waltham, MA) was diluted 1:500 in 1% (w/v) Casein/PBS solution and 30 μL added to the well. The plate was then incubated for 10 min at room temperature before a PBS-T wash.

Following the removal of the secondary antibody, a Poly80 Horseradish peroxidase enzyme is prepared at 1:5 dilution ratio in 1% (w/v) Casein/PBS solution and incubated at 10 minutes at room temperature prior to another PBS-T wash.

### Measurement of Electrochemical Signal and Data Analysis

Sixty microliters of 1 Step Ultra-TMB (ThermoFisher, Waltham, MA) is added to the wells and the plate immediately inserted into the electrochemical reader device, with a fixed potential of -200 mV and simultaneous measurement of electrochemical current for all 96 wells is measured 2 separate times over a 60 second period.

Signal of the last 10 seconds of the readout procedure is averaged for final quantitative signal value. All saliva samples tested were normalized by subtracting the signal of the polymer only wells with the antigen-polymer coated wells. Standards were also compared to calibrators for quantification.

### Human Subjects

Research protocol and consents were approved by the Western Internal Review Board (Study #1302611, Expiration Date: March 19^th^, 2022).

Individuals under the age of 18 years and individuals receiving immunosuppressive drugs or cancer chemotherapy were excluded from the study.

Volunteers who had previously received a Pfizer (BioNTech), Moderna, or Johnson and Johnson Vaccine for SARS-CoV-2 were consented. Subjects were issued a questionnaire collecting information about vaccination dates and vaccine type, along with questions to eliminate subjects who were immunocompromised or taking immunosuppressants.

The study comprised of a longitudinal and a cross-sectional study cohort. For the longitudinal study cohort, a cohort receiving either the Pfizer (n=15) or Moderna (n=27) mRNA vaccine were monitored with a first-morning twice-weekly collection. Collections lasted for as long 8 months post vaccination for some individuals. We analyzed saliva at a single time point for another 31 and 24 individuals receiving the Pfizer and Moderna vaccines respectively. This has allowed us to make several conclusions regarding the kinetics of COVID-19 vaccine response in community vaccinated populations. In all more than 840 samples have been collected and analyzed.

### Sample Processing Device and Protocol

Saliva samples were collected using the Orasure^®^ Oral Fluid Collection Device (Orasure Technologies, Bethlehem, PA), which consists of an absorbent pad on the end of a long wand and a collection tube with preservative solution. Subjects insert the absorbent pad in the mouth for a minimum of 2 minutes in order to absorb adequate saliva fluids. The absorbent pad is then immersed into a collection tube and the wand broken at a scored breakpoint to allow the device to be securely capped. Individuals participating in longitudinal studies placed the capped collectors in a zip lock bag and then into their home freezer until shipping them to the laboratory at ambient temperature. Individuals providing single samples kept the samples at room temperature until shipping to the laboratory.

Upon receipt at a central laboratory, the tube is uncapped, the pad gently pressed on the inside of the tube to squeeze out saliva, and the saliva transferred into a labeled microcentrifuge tube for testing. Samples are stored at -80C for long term storage. Previous evaluations of the collection system *(17)* have demonstrated the system can store the samples at room temperature for greater than 10 days without significant degradation in antibody levels.

### Variant Antigen (Delta)

To determine whether the antibodies of vaccinated or convalescent patients were reactive to the delta variant, we used SARS-CoV-2 variant S1 Antigen B.1.6.617.2 (40591-V08H23 SinoBiological, Wayne, PA), a recombinant antigen that included T194, G142D, E156G, 157-158 deletion, and the L452R, T478K, D614G, 681R mutations. Identical amounts of this variant antigen are immobilized in the gel and the standard assay is performed.

## RESULTS

### Kinetic Studies

We analyzed 42 patients vaccinated patients (27 Moderna, 15 Pfizer) who provided twice-weekly samples for a period of several weeks. The general patterns for all patients were comparable to one another. Figure 1 demonstrates 8 representative samples from this longitudinal. The curves are oriented with zero time being the date of the second injection. The general patterns are similar for individuals receiving both vaccines with a spike in antibody production 1-2 weeks following the second injection followed by a steady decrease in antibody levels. There were, however, differences in the robustness of response, with most volunteers having robust responses with 100 ng / mL – 200 ng / mL of IgG as a peak response followed by a gradual decrease in levels over time.

**Figure 1.**
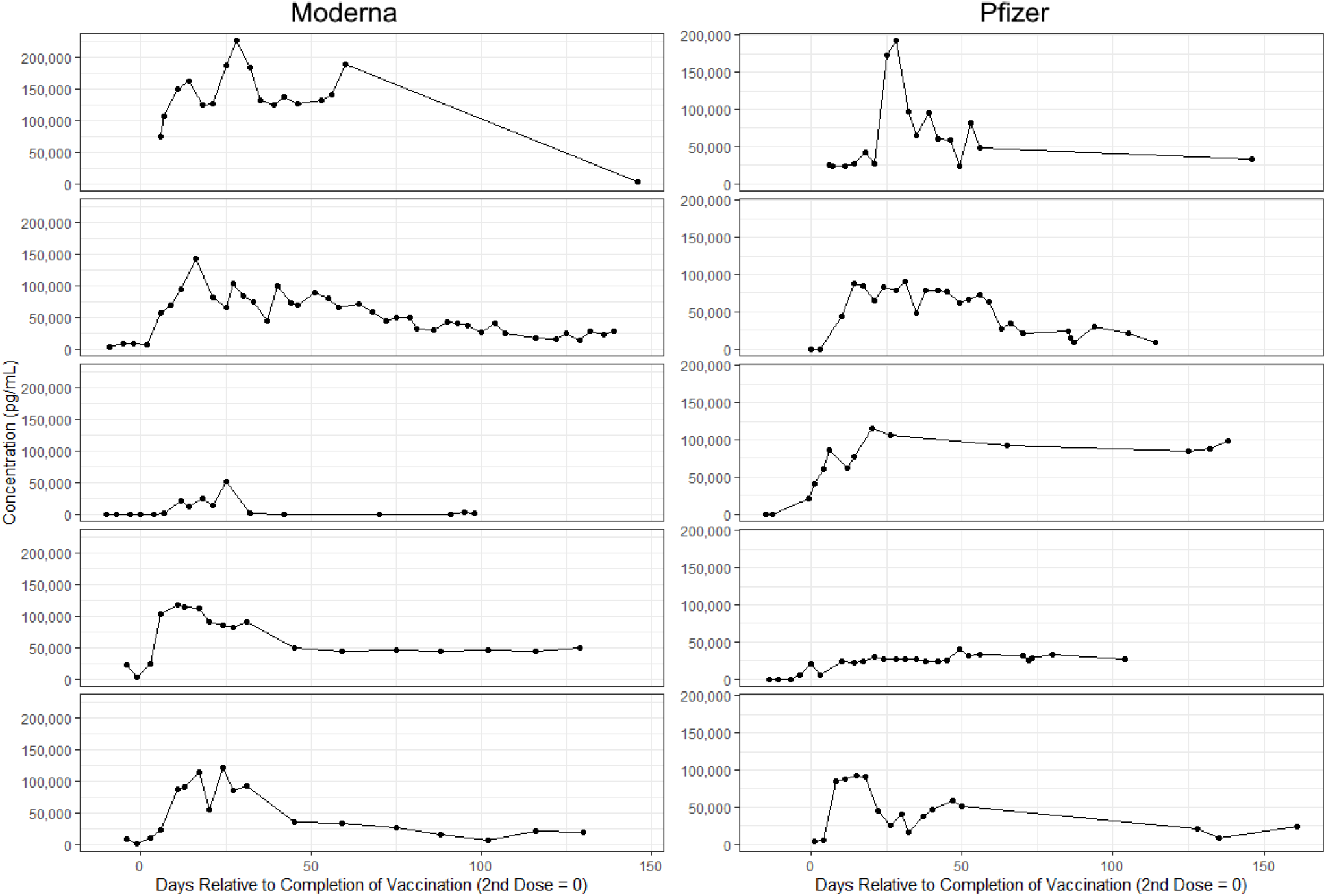
Representative Individual Kinetic Experiments: Pfizer and Moderna, with graphs centered around time 0 being the day of the second vaccination.

As can be seen in Fig 1, however, 2 individuals responded with a maximum level of 50 ng / ml. Individual 3 in the Pfizer group had a peak response of approximately 50 ng / ml and then stabilized at approximately 25 ng / ml. Individual 3 in the Moderna group had a short-duration peak of 50 ng /ml followed by a return to baseline 30 days post second vaccination. All but 2 patients experienced a gradual, but steady decline in antibody levels. These decreasing levels may correlate with the need for booster vaccinations.

Clinical trials data revealed an approximate 50% protection for individuals 2 weeks after having received their first immunization with either the Modern or Pfizer vaccine *(1-5)*. We wondered whether this could be a function of antibody response in vaccinated patients. Of the 42 subjects that were serially monitored, 36 supplied samples before the second dose. In the subjects that were collected prior to the second dose, 88% of the Moderna subjects had detectable antibodies prior to the second dose and 50% of the Pfizer subjects had detectable antibodies prior to the second dose (see Table 2).

### Summary Statistics for Kinetic Studies

Table 1 shows the summary statistics for the volunteers participating in the kinetic studies. The average time to maximum antibody level was 22 days for Moderna and 30 days for Pfizer. The maximum levels were nearly identical for the 2 vaccines with Moderna vaccinated individuals having average peak levels of 127 ng / ml and 130 ng /ml respectively for Moderna vaccinated and Pfizer vaccinated individuals. These levels are similar to those we observed in convalescent hospitalized COVID-19 patients and 5 fold higher than more mildly symptomatic individuals (17).

**Table 1:**
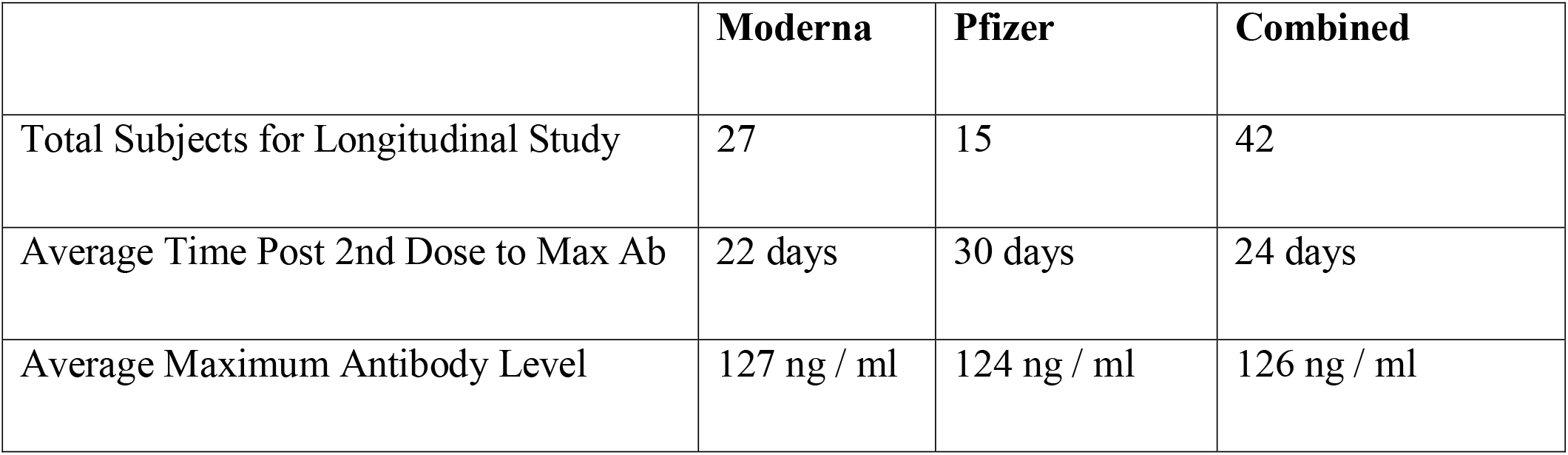
Summary Statistics for kinetic studies on vaccinated volunteers

|  | **Moderna** | **Pfizer** | **Combined** |
| --- | --- | --- | --- |
| Total Subjects for Longitudinal Study | 27 | 15 | 42 |
| Average Time Post 2nd Dose to Max Ab | 22 days | 30 days | 24 days |
| Average Maximum Antibody Level | 127 ng / ml | 124 ng / ml | 126 ng / ml |

In addition to the 42 volunteers participating in the kinetic studies we had an additional 53 individuals submitted single samples for this study. Figure 2 is a summary of all the data representing 840 individual time points including the multiple time points for the 42 volunteers submitting multiple samples and the 53 volunteers submitting only one samples. The samples are normalized to days after receiving the second vaccine dose.

**Figure 2.**
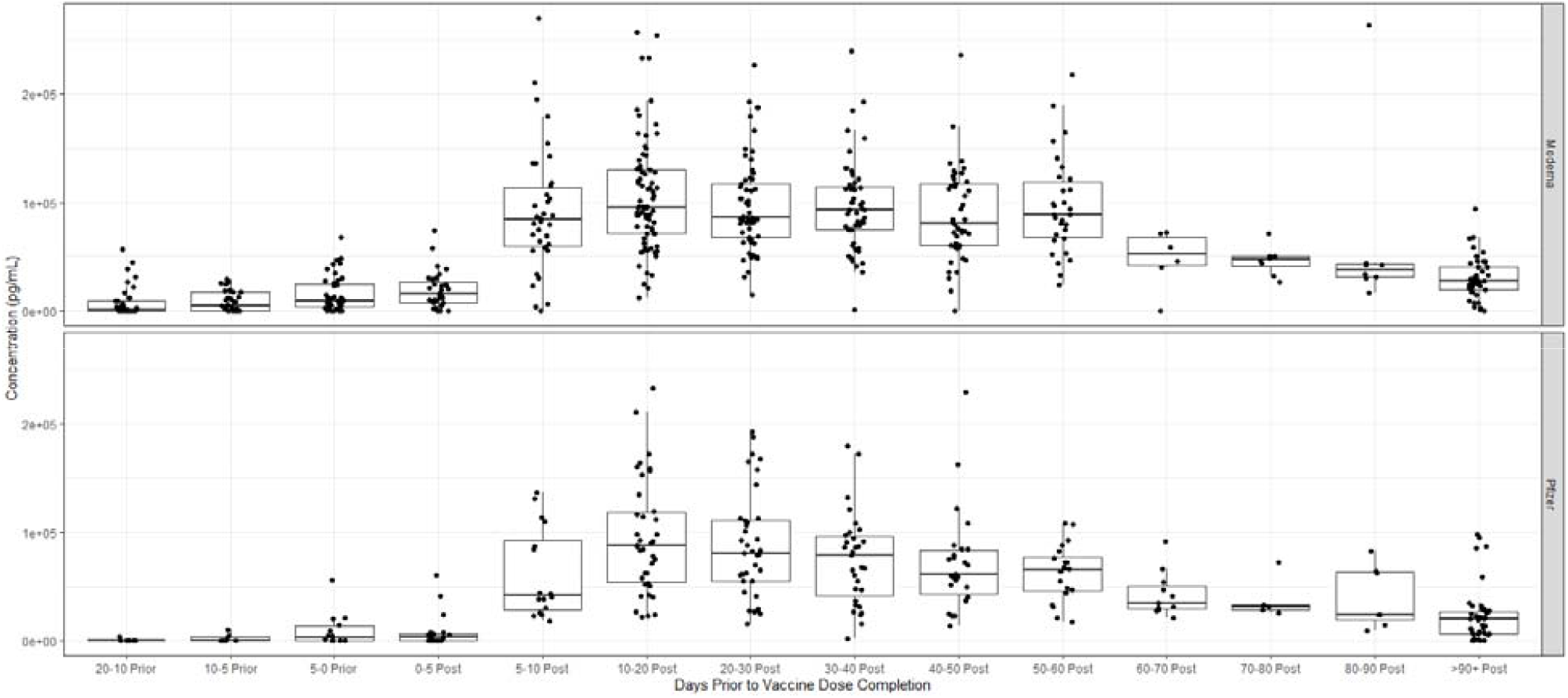
Samples collected from volunteer subjects (n=99) at different time intervals for Pfizer (n=47) and Moderna (n=52) vaccines were tested and binned to different time intervals relative to completion of second dose of mRNA vaccine.

Figure 2 is a summary of these data showing a box plot of weekly antibody levels for individuals both pre and post second vaccination. The trend is clear. Robust antibody levels are present for the initial 60 days following the second vaccination. Subsequently, levels begin falling gradually, but consistently. The data demonstrate a steady decrease in antibody levels with increasing time following vaccination.

### Summary of subjects with significant drops from peak value

Table 3 is a summary of volunteers who have experienced drops of more than 90%, 95%, and 99% from their peak values grouped by vaccine type. Although a higher percentage of Pfizer vaccinated patients experience a decrease of 90% or more (33% vs 15%) and 95% or more (13% v 8%) no Pfizer volunteers experienced a 99% drop whereas one Moderna vaccinated patient experienced a 99% drop in antibody levels (see Table 3). The numbers are not sufficient to form any conclusions regarding any potential differences between the 2 vaccines but do show that, with time, antibody levels drop to 90% of their peak level in 20% of community vaccinated individuals.

**Table 2.** Summary of subjects with measurable antibodies prior to completion of second dose

|  | **Moderna** | **Pfizer** | **Combined** |
| --- | --- | --- | --- |
| Individuals With Data Collected Prior to 2nd Dose | 26 | 10 | 36 |
| Antibody Produced Before 2nd Dose | 23 (88%) | 5 (50%) | 28 (77%) |

**Table 3:**
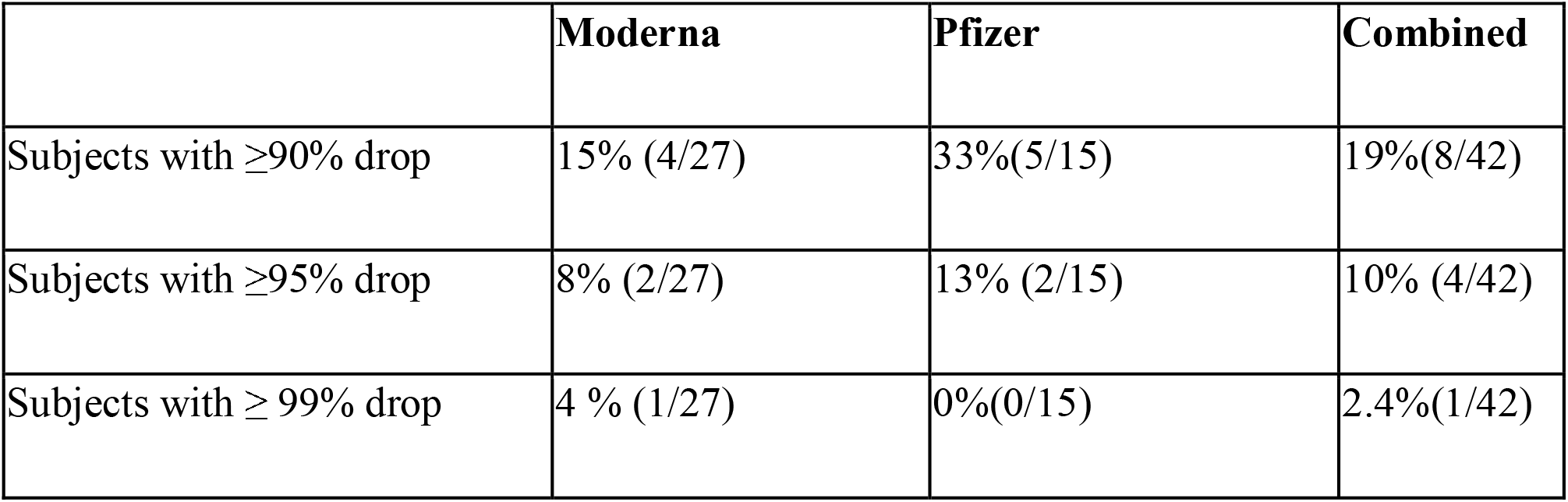
Individuals with large drops in antibody levels

Figure 3 are the kinetic plots of the 9 vaccinated individuals who experienced >90% drops in antibody levels following vaccination. It is apparent that that there is no correlation with the original peak value with prediction of eventual 90% drop in antibody level. Two individuals had peak levels above 200 ng / ml indicating a robust initial response. There were two volunteers who had initial peak values of only 50 ng / ml who also dropped to low levels. Two individuals (5%) dropped to undetectable levels; one a Pfizer patient and one a Moderna patient.

**Figure 3.**
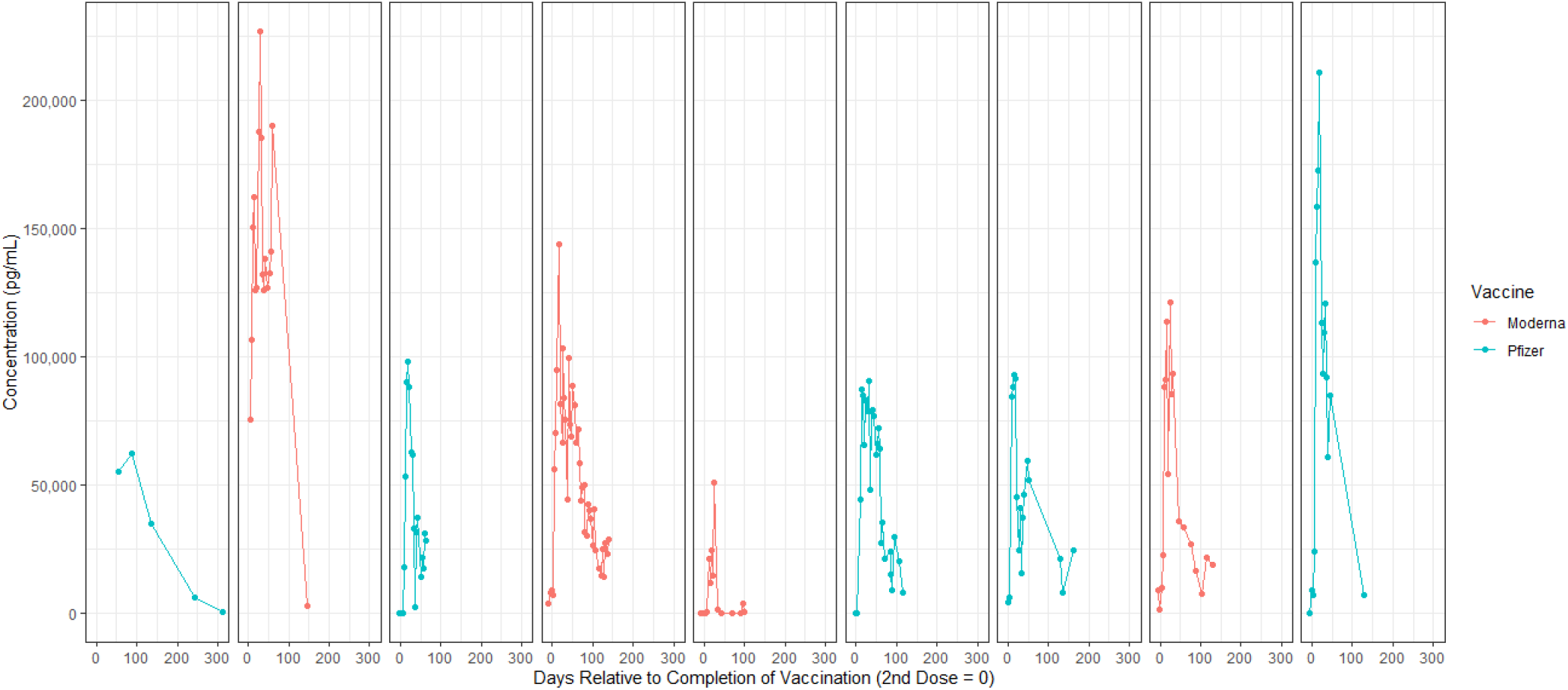
Plot of individuals measured with over 90% drop from peak.

### Case Study: Prednisone Effects

A male patient in their sixties was given a 3 week course of prednisone (50 mg / day for 14 days followed by a week of tapering) for a nasal polyp approximately 2 months after his second dose of vaccine. The kinetics of his antibody production is shown in Figure 4 with the time period that the prednisone was being administered is highlighted in grey. As can be seen, antibody levels began falling with the onset of treatment to baseline levels and remained suppressed for several weeks following the taper. However, antibody levels did rebound and then began to slowly decline thereafter.

**Figure 4.**
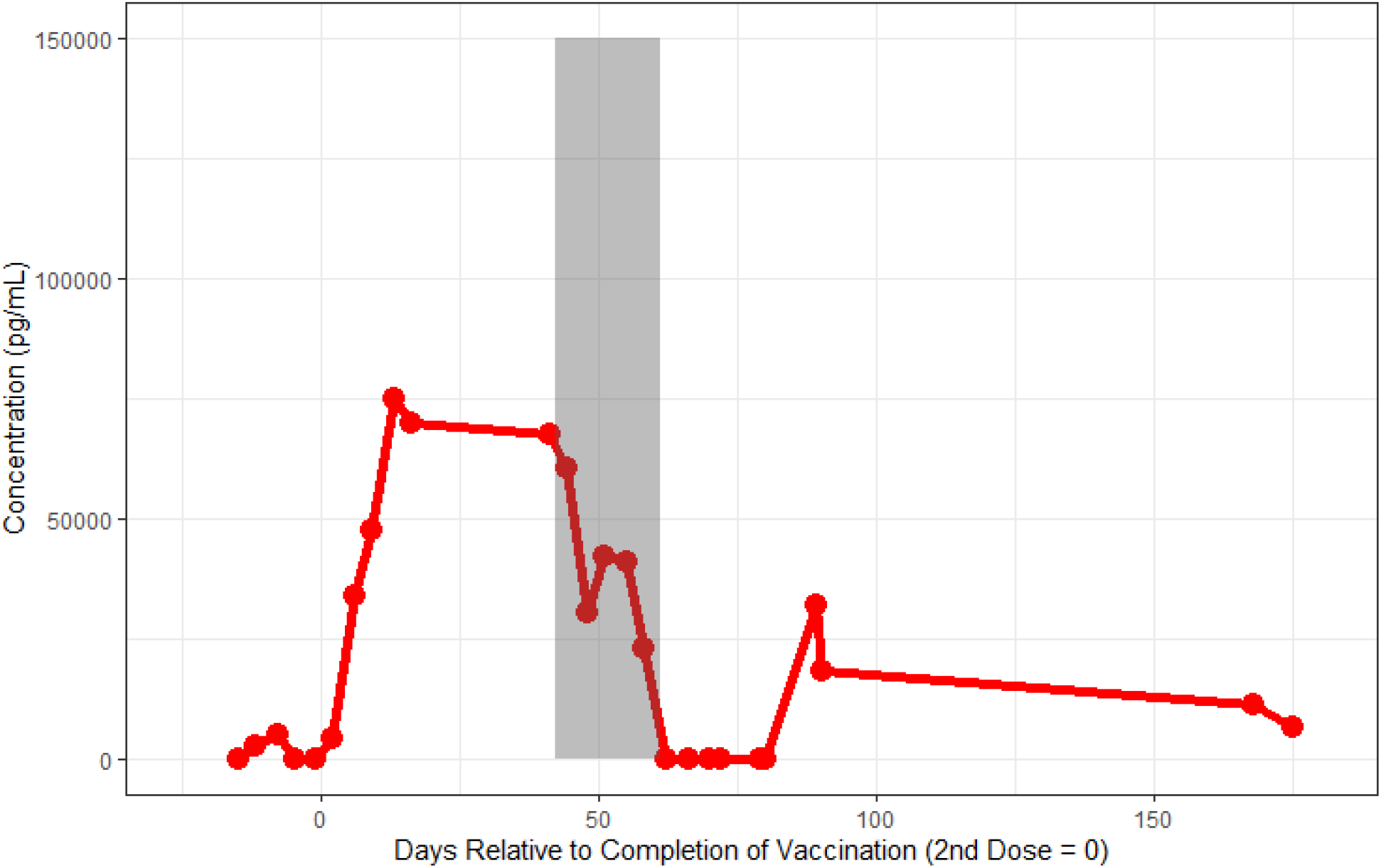
Case Study Plot of Pfizer vaccinated individual who was administered prednisone following his vaccination. Shaded area of grey indicates the period of time where prednisone was taken.

### Evaluation of Delta Variant Antigen to Salivary Antibodies

The Delta variant of SARS-CoV-2 has become the predominant variant in the United States. We therefore investigated whether antibodies present in convalescent and vaccinated patients are capable of recognizing the S1 antigen of the Delta variant. We designed an Amperial^®^ assay substituting monoclonal S1 delta variant antigen for the wild type S1 antigen (see Methods). The standard curves for this assay compared to the wild type antigen assay are shown in Figure 5A and demonstrate very similar assay characteristics. There is some indication of slightly reduced binding efficiency for monoclonal Anti-S1 antibodies to the Delta variant versus the wild type S1 antigen but these differences are not enough to alter testing results.

**Figure 5A:**
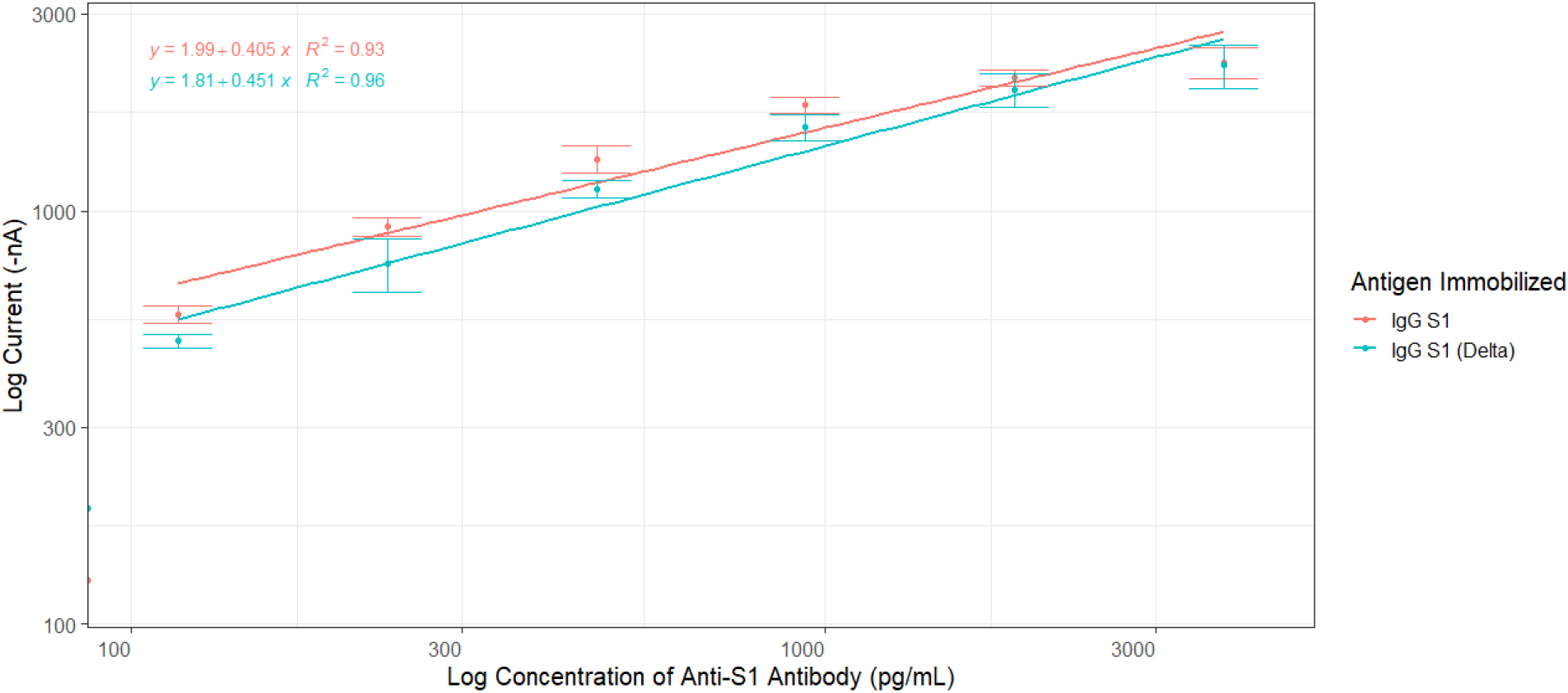
Standard curves for both the Wild type and Delta Variant S1 antigens

Next, we investigated whether antibodies present in convalescent individuals and vaccinated individuals could recognize and bind to the Delta variant S1 antigen. A total of 3 pre 2019 samples were used as controls, with also 1 immunodeficient organ transplant patient run for reference. Three samples from convalescent patients with detectable antibody levels were used. These patients were infected before the delta variant emerged. In addition, 3 Pfizer and 4 Moderna vaccinated individuals with detectable antibody were analyzed in parallel by both the Wild Type assay and the Delta Amperial^®^ assay.

These data are shown in Figure 5B. For all cases of vaccinated and convalescent subjects there was no significant reduction of apparent antibody concentration in saliva to the Delta variant versus the Wild Type. These data demonstrate that antibodies made against the current Moderna and Pfizer vaccines do recognize the S1 domain of Delta variant Spike Protein. In addition, individuals infected prior to the emergence of the Delta variant Convalescent also developed antibodies that recognize the Delta variant. Although this cannot insure an equal protection level against serious infection, it is reassuring.

**Figure 5B:**
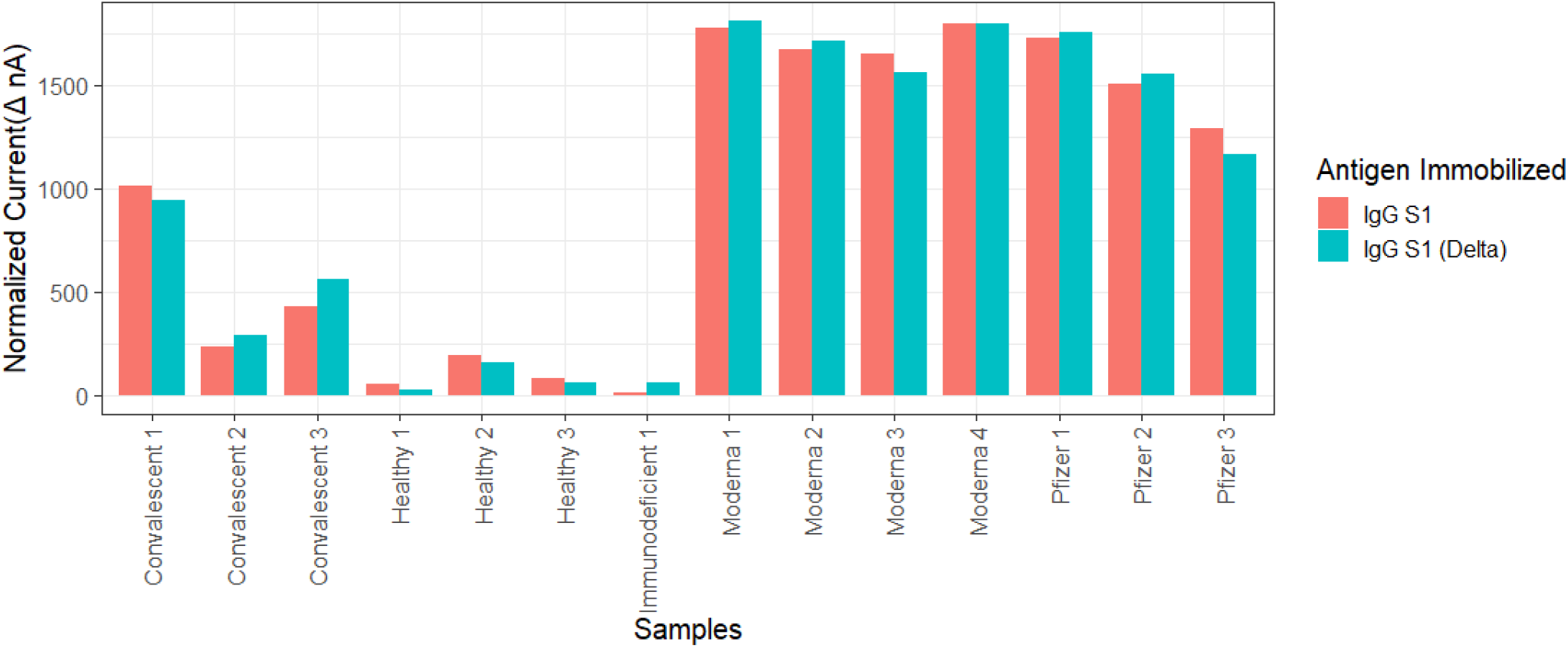
Comparison of Wildtype Anti SARS-CoV-2 IgG S1 and B.1.6.617.2 variant S1 SARS-CoV-2 antigen

## DISCUSSION

This study demonstrates that, although all individuals vaccinated with Pfizer or Moderna vaccine develop a robust antibody response, the response wanes over time. Approximately 20% of vaccinated individuals experience a drop off of >90% after 90 days post vaccination. In 2 (5%) of serially monitored patients, antibody levels became undetectable. The ability to monitor vaccine response non-invasively can be an important way to identify individuals who may require additional injections without straining stretched health care resources.

Although some individual variability is seen among individuals in terms of fluctuating levels, it is easy to determine trends over time using serial saliva monitoring. Previous studies have determined that serum and saliva levels are highly correlated *(17-19)*. However, one cannot predict the absolute serum level by measuring the salivary level. It appears that each individual has his or her own gradient between saliva and serum. However, as the data in the article demonstrate, that gradient remains relatively constant over time allowing longitudinal monitoring to be performed.

Herd immunity from widespread community vaccination is a key component in preventing COVID-19 infections and in curbing the pandemic. Several questions remain unanswered. Our data can help provide the answers to some of these questions:

Does everyone respond to vaccination with a robust immune response? In this study all vaccinated individuals did respond, although some with much lower antibody levels than the average. Antibody monitoring post vaccination could identify the individuals who did not react to vaccination with a robust antibody response and allow these individuals to have an immunologic evaluation or an additional injection or a different vaccine type.

Will booster vaccination be necessary? Recent data regarding breakthrough infections and CDC recommendations are for immunocompromised individual and patients receiving immunosuppressive therapy should receive a third dose of the vaccine regardless of timing. Health care workers and high-risk individuals are scheduled to receive boosters in September. Our data supports this approach in that most individuals experience a continuous drop in antibody levels with time and 5% of individuals dropped to undetectable levels. Although it is not clear what level of antibody, if any, is necessary to prevent COVID-19 infection, individuals with baseline levels of antibodies may be at higher risk to acquire infection.

Will a fourth vaccination be needed? Future kinetic studies will be necessary to determine if antibody levels will remain stable following a third vaccination. Noninvasive monitoring using saliva home collection provides a low cost, effective way to perform population monitoring of vaccine levels following a third vaccination.

Will the current vaccines protect against the Delta variant? Our data shows that antibodies produced in convalescent patients or mRNA vaccinated subjects do recognize the Delta variant. Although this cannot insure an equal protection level against serious infection it is reassuring.

Could any individuals in low-risk groups benefit from booster vaccination? Our data suggests that about 20% of individuals experience a fall of >90% of antibody levels 3 months following completion of their vaccination protocol. These individuals might benefit from early booster shots to prevent breakthrough infections. If an individual with a low antibody level is identified by saliva testing, further evaluation can be performed using serum titers to confirm the initial observation.

We should stress that it has not been determined what level of circulating IgG antibody, if any, is necessary to prevent COVID infection. The data in this article must be interpreted in that light. The ability to noninvasively and cost efficiently quantitates COVID-19 antibody levels could be an important tool in investigating the relationship between circulating antibodies to immunity.

The results presented in this work regarding the salivary monitoring of SARS-CoV-2 are congruent with recommendations given by the USA CDC and established literature regarding SARS-CoV-2 antibody in vaccinated populations. While there still remains a need for a more comprehensive evaluation of the relationship between salivary SARS-CoV-2 antibodies and those present in the blood, our work demonstrates that our noninvasive quantitative saliva assay could be valuable for evaluating a community vaccinated population and to further investigate the relationship between circulating antibody to COVID-19 immunity.

## Data Availability

Data will be available on demand

## Acknowledgments

The authors wish to thank our volunteers who participated in the study. Funding sources: U18 TR003778, U54 HL119893 (to DTWW)

## Disclaimers

D. Wong is consultant to Colgate-Palmolive, Mars Wrigley and GSK. D. Wong also have equity in Liquid Diagnostics and RNAmeTRIX. DW, CS, MT, RB are shareholders in Liquid Diagnostics LLC. MT is a paid consultant of Liquid Diagnostics LLC. RB is a consultant to Amgen, Bristol Myers Squibb.

## Author Bio

(first author only, unless there are only 2 authors)

Michael Tu, PhD is the Chief Scientific Officer of Liquid Diagnostics LLC. His training and background in biomedical engineering and biosensors, and his work primarily emphasizes the development of novel diagnostic tools for disease related biomarkers.

